# Comparison of large language models for citation screening: A protocol for a prospective study

**DOI:** 10.1101/2024.06.26.24309513

**Authors:** Takehiko Oami, Yohei Okada, Taka-aki Nakada

## Abstract

**Background:** Systematic reviews require labor-intensive and time-consuming processes. Large language models (LLMs) have been recognized as promising tools for citation screening; however, the performance of LLMs in screening citations remained to be determined yet. This study aims to evaluate the potential of three leading LLMs - GPT-4o, Gemini 1.5 Pro, and Claude 3.5 Sonnet for literature screening.

**Methods:** We will conduct a prospective study comparing the accuracy, efficiency, and cost of literature citation screening using the three LLMs. Each model will perform literature searches for predetermined clinical questions from the Japanese Clinical Practice Guidelines for Management of Sepsis and Septic Shock (J-SSCG). We will measure and compare the time required for citation screening using each method. The sensitivity and specificity of the results from the conventional approach and each LLM-assisted process will be calculated and compared. Additionally, we will assess the total time spent and associated costs for each method to evaluate workload reduction and economic efficiency.

**Trial registration:** This research is submitted with the University hospital medical information network clinical trial registry (UMIN-CTR) [UMIN000054783].

## Background

A systematic review comprises several steps, including the formulation of a query, citation screening, qualitative assessment, and meta-analysis. Among these processes, citation screening is known to be time-consuming and resource-intensive [1-3]. Although recent studies have explored machine learning applications for citation screening [4-9], achieving both time efficiency and high accuracy continues to be challenging [9-11].

The advent of large language models (LLMs) has illuminated new possibilities in natural language processing and the completion of complex tasks [12, 13]. These tools have demonstrated potential in revolutionizing citation screening through their sophisticated comprehension and human-like response generation capabilities [14, 15]. Prior research has suggested the potential of LLMs in citation screening tasks [16]. However, comprehensive studies comparing the performance of LLMs are lacking.

Therefore, we will seek to investigate the performance of different LLMs in screening citations. This study aims to evaluate and compare three recent LLMs— GPT-4o, Gemini 1.5 Pro, and Claude 3.5 Sonnet—in their ability to conduct citation screening.

## Methods

### Study design and settings

We will conduct a prospective study to evaluate the performance of LLMs in citation screening. To enhance the transparency and accessibility of our methodology, we have submitted our comprehensive review protocol to the medRxiv pre-print platform. Additionally, we have registered our study with the University Hospital Medical Information Network (UMIN) clinical trials registry (UMIN000054783).

### Clinical questions in the J-SSCG

Our study will evaluate the accuracy of LLMs using clinical questions (CQs) from the upcoming J-SSCG 2024, an updated version of the 2020 guidelines. Developed by the Japanese Society of Intensive Care Medicine (JSICM) and the Japanese Association for Acute Medicine (JAAM), these guidelines specifically address sepsis and septic shock management in Japanese clinical settings [17].

We will employ the same five clinical questions (CQs) as in our previous research (Table 1) [11]. These CQs underwent comprehensive literature reviews across multiple databases, including CENTRAL, PubMed, and Ichushi-Web. The working group meticulously developed search strategies to guarantee the inclusion of all relevant studies. Our search was confined to literature in Japanese and English. For J-SSCG 2024, we utilized EndNote as our citation management tool. This software facilitated the downloading, compiling, and removal of duplicates from all titles and abstracts gathered during our literature search.

**Table 1.** The list of the patient/population/problem, intervention, and comparison of the selected clinical questions.

|  | Patient, population, problem | Intervention | Comparison |
| --- | --- | --- | --- |
| CQ1 | Adult patients (18 years old or older) diagnosed with or suspected of having infection, bacteremia, or sepsis | Balanced crystalloid administration | 0.9% sodium chloride administration |
| CQ2 | Adult patients (18 years old or older) with sepsis, or suspected as sepsis, infection, bacteremia or patients admitted to ICU | Targeting a higher mean arterial pressure | Targeting a lower mean arterial pressure |
| CQ3 | Adult patients (18 years old or older) with sepsis presenting with severe metabolic acidosis or patients admitted to ICU | Sodium bicarbonate administration | No sodium bicarbonate administration |
| CQ4 | Adult patients (18 years old or older) with sepsis or septic shock | Usual care with at least one of the following tissue perfusion parameters: lactate/lactate clearance, capillary refill time, ScvO<sub>2</sub>/SvO<sub>2</sub>, and P(v-a) CO<sub>2</sub>/C (a-v) O<sub>2</sub>. | Usual care with different parameters mentioned in the interventional group, or standard care without the utilization of any specific tissue perfusion parameters |
| CQ5 | Adult patients (18 years old or older) with sepsis, sepsis-induced hypotension, or septic shock | Restrictive fluid management, which aims to reduce the amount of fluid therapy for up to 24 h | Conventional fluid management or non-restrictive fluid management defined by authors |

CQ: clinical question; ICU: intensive care unit

### Conventional citation screening

Members of J-SSCG 2024 transferred files processed in EndNote to Rayyan, a software specifically designed to facilitate systematic reviews. The screening protocol involved two independent reviewers each assessing the title and abstract of each study. Disagreements were resolved through collaborative discussions or, when necessary, by consulting a third reviewer for an impartial evaluation. As a standard reference for assessing accuracy, we will utilize the screening results from conventional citation screening methods.

### Large language model

Our prospective study will critically assess the accuracy, time efficiency, and cost of three LLMs, including GPT-4o (OpenAI, San Francisco, CA), Gemini 1.5 Pro (Google, Mountain View, CA), and Claude 3.5 Sonnet (Anthropic, San Francisco, CA), released on May 13, 2024, May 23, 2024, and June 21, 2024, respectively. After importing the dataset from citation managers using the same procedure as the conventional tool for citation screening, we interfaced the dataset with the Application Programming Interface (API) using pandas (version 1.0.5) in Python (version 3.9.0). We will utilize the publicly available API for each LLM. To conduct LLM-assisted citation screening, we developed a command prompt that enables the LLMs to automatically execute the citation screening process. For each query, we will adhere strictly to the same phrases outlined in the framework of CQs that the J-SSCG2024 members formulated for conventional citation screening.

#### Prompt

You are conducting a systematic review and meta-analysis, focusing on a specific area of medical research. Your task is to evaluate research studies and determine whether they should be included in your review. To do this, each study must meet the following criteria:

Target Patients: ------------

Intervention: ----------

Comparison: ---------

Study Design: The study must be a randomized controlled trial.

Additionally, any study protocol that meets these criteria should also be included.

However, you should exclude studies in the following cases:

The study does not meet all of the above eligibility criteria.

The study’s design is not a randomized controlled trial. Examples of unacceptable designs include case reports, observational studies, systematic reviews, review articles, animal experiments, letters to editors, and textbooks.

After reading the title and abstract of a study, you will decide whether to include or exclude it based on these criteria. Let’s think step by step. Please answer with include or exclude only.

Title: ---------

Abstract

---------------------------------------------

Through the process of the automated citation screening using LLMs, inclusion or exclusion decisions was provided without prior context. Upon completion of this phase, we will review the judgement documented in the output file. The source code for this procedure will be made available in a public GitHub repository (https://github.com/seveneleven711thanks39/gpt-assisted_citation_screening.git).

### Data collection

This study will collect and evaluate the following variables:

#### Accuracy

Accuracy: After compiling the number of references included by each LLM, we will compare the sensitivity and specificity of these results to those obtained through manual screening.

#### Time Efficiency

The time required for citation screening with each LLM will be measured and compared to that of manual methods.

#### Cost

The study will assess the overall costs associated with API usage, based on a usage-based billing system.

### Statistical analysis

To assess and compare the accuracy of LLMs, we will calculate the sensitivity and specificity of citations accurately identified as “relevant” by the LLMs. Our primary analysis will utilize the results from the qualitative assessment of conventional screening as the standard reference. The secondary analysis will employ the results from the title and abstract review of conventional screening as the standard reference.

To assess time efficiency, we will aggregate the durations of systematic review sessions across all clinical questions. To calculate the cost of LLM-assisted citation screening using APIs, we will document the total charges incurred under the pay-as-you-go system. Additionally, we will perform a sensitivity analysis to investigate how variations in the LLM’s prompts influence screening accuracy, focusing on the effects of prompt engineering on the model’s performance in citation assessment tasks. In our analysis, we will present continuous data as means and standard deviations or medians and interquartile ranges, depending on the distribution of the data. For the statistical analysis, we will use GraphPad Prism 10 (GraphPad Software, San Diego, CA).

## Data Availability

All data produced in the present study are available upon reasonable request to the authors.

